# Impact of Skin Tone, Environmental, and Technical Factors on Thermal Imaging

**DOI:** 10.1101/2025.05.08.25327244

**Authors:** Sharon Eve Sonenblum, Kathleen Jordan, Glory Tomi John, Andrew Chung, Miriam Asare-Baiden, Jordan Pelkmans, Judy Wawira Gichoya, Vicki Stover Hertzberg, Joyce C. Ho

## Abstract

**Background:** Erythema, an early visual indicator of tissue damage preceding pressure injuries (PrIs), presents as redness in light skin tones but is harder to detect in dark skin tones. While thermography shows promise for early PrI detection, validation across different skin tones remains limited. Furthermore, most protocols and models have been developed under highly controlled conditions.

**Objective:** To evaluate how environmental and technical factors (i.e., patient positioning, lighting, distance, camera type) and skin tone impact thermal imaging measurements and temperature change.

**Methods:** This pre-post experimental study enrolled 35 healthy adults (30 with Monk Skin Tone Scale ≥6). Melanin Index was measured on the volar forearm using the SkinColorCatch. After baseline imaging, a 15.5°C cooling stone was placed on one posterior superior iliac spine (PSIS) for 5 minutes. Thermal images were then collected with either the FLIR E8-XT or the FLIR ProOne camera under varied conditions: two lighting types (overhead room versus localized LED ring light), three postures (side-lying, side-lying with forward knee placement, and side-lying with rearward knee placement), and two camera-to-body distances (35cm and 50cm from the PSIS). The cooling/imaging procedure was repeated using the alternate camera, and data were analyzed using mixed-methods model.

**Results:** Temperature change was effectively detected across all skin tones, with cooling resulting in a -3.7 ± 1.2°C difference between the region of interest (ROI) and control region. Camera type significantly affected measurements, with the ProOne recording 1.04°C less cooling than the E8-XT. Distance had minimal impact (0.11°C cooler at 50cm vs 35cm at baseline), with no significant difference when comparing ROI to control regions, while lighting and posture had no impact on measurements. Skin tone influenced cooling measurement, with higher melanin levels showing greater temperature changes. A 0.98°C difference was observed between the lightest and darkest skin tone groups.

**Discussion:** Our findings confirm thermal imaging’s robustness across varied environments, with the minor distance effects mitigated through perpendicular measurements and relative temperature comparisons. Significant discrepancies between thermal cameras (>1°C) highlight that these technologies cannot be used interchangeably when establishing thresholds. While effective across all skin tones, the observed differences in cooling response suggest tailored thresholds may be necessary for darker skin tones. Future research should focus on clinical validation across diverse populations to enhance PrI detection accuracy.

## Introduction

Thermal imaging measures heat radiating from the body and is a tool with many potential clinical applications.

Of particular interest is the use of thermography for early detection of pressure injuries (PrIs), which has shown promise. When incorporated in a prevention bundle, thermography may also reduce the rate of PrI development. (1–9) Thermal imaging has also been used to detect acute appendicitis (10), deep vein thromboses (11), coccyx pain (12), myofascial trigger points (13), hypoperfusion in critically ill patients (14), and wound healing (15,16), among other applications. Despite widespread application, studies do not report the skin tone of participants and when available, participants with very dark skin tones were not studied.

Erythema, characterized by redness in persons with light skin tones, is an early visual indication of tissue damage that often precedes a PrI. In individuals with dark skin tones, however, erythema often presents as hyperpigmentation, which is more difficult to detect. Difficulty detecting skin color changes in persons with dark skin tones contributes to underlying health disparities related to PrIs, such as discovery at later stages and slower healing among Blacks, Hispanics, Native Americans, and Asians. (17–19) Without early detection, individuals with dark skin tones suffer disproportionately, enduring longer hospital stays, have more severe infections, reduced wellbeing, and in some cases, premature death. (17,18,20–22) Thus, it has been suggested that non-invasive bedside accessible technologies are needed to identify PrIs to reduce health disparities due to skin color. (23–26). In fact, thermography was highlighted in the 2019 International Pressure Injury Prevention and Treatment Clinical Practice Guideline as a priority for future research. (27)

However, as mentioned previously, limited work has been done to validate the use of thermal imaging across different skin tones. Emissivity of human skin does not vary according to skin pigmentation, meaning that the thermal camera’s settings do not need to be changed according to the patient’s skin tone, a finding that is promising for the potential of thermography to be effective across diverse skin tones. (28) However, many other environmental, camera, and subject related factors may contribute to the accuracy of thermal measurements. (29,30) Furthermore, in a recent review on thermography’s accuracy in PrI detection, only one out of the eight included papers considered skin tone. (8) That study acknowledged the technology’s effectiveness on Fitzpatrick Skin Types I to III, but identified a need for improved approaches in darker skin tones. (31) A recent clinical trial of a thermography system in a long-term care setting did not report the demographics of the population nor the efficacy on the patients with dark skin, but the city in which the hospital was located included approximately 10% African American residents. (9)

There have been several devices that, when used unvalidated in individuals with dark skin, have turned out to have inadequate performance resulting in real world consequences. For example, over-estimation of oxygenation with pulse-oximetry led to delays in treatment for African American patients, (32,33) while infrared thermometers under-reported fevers in African Americans admitted to the hospital. (34)

Furthermore, guidelines for using thermography dictate highly controlled conditions, specifying everything from the camera distance and room lighting to the floor surface and HVAC conditions. (35,36) Most studies designed to develop a protocol for detecting erythema similarly use highly controlled conditions. (5,7) Clinical environments are not nearly that controlled, resulting in challenges when implementing models developed in controlled environments into the real world. Therefore, the objective of this study was to evaluate how environmental and technical factors (i.e., patient positioning, lighting, distance, camera type) and skin tone impact thermal imaging measurements and temperature change.

## Materials and Methods

This study used a pre-post experimental design with multiple instances of induced skin cooling on the lower backs of healthy adults.

### Participant Population

This study enrolled 35 healthy adults between March 26^th^ and June 25^rd^ of 2024, 30 of whom had dark skin tones, defined as Monk Skin Tone (MST) Scale level 6 or greater when measured at the inner forearm, and 5 who had skin tones with MST level 5 or lower. (37) Participants had to be able to consent and were excluded if they did not speak English, were a member of a special population including women identifying as pregnant, individuals with a diagnosed communicable skin disease, individuals with a skin disease that might have been irritated by inducing erythema, or a bleeding disorder that made the individual prone to bruise more easily. This study was approved by Emory University’s Institutional Review Board (eIRB number 00005999).

### Instrumentation and Measurements

#### Demographic Data

Participants completed an electronic REDCap survey containing questions about demographic data and information about height, weight, and smoking status.

#### Colorimetry

The SkinColorCatch® (Delfin Technologies Ltd, Kuopio, Finland), a digital colorimeter, was used to describe the Melanin Index of the participant’s forearm. The SkinColorCatch® measures reflected light from the skin with a red, green, blue (RGB) color sensor. The SkinColorCatch® calculates a melanin index based on changes in the red-green light absorption, which are impacted by hemoglobin and melanin levels in the skin. (38,39) Skin tone at the inner forearm was classified using the modified Eumelanin Human Skin Colour Scale (Eumelanin Scale-Modified) (40,41), adapted from the scale initially described by Dadzie, et al. (42). The melanin indices for the modified Eumelanin Human Skin Colour Scale were defined as Eumelanin low: <25, Eumelanin intermediate low: 25 to <37.5, Eumelanin intermediate: 37.5 to <50, Eumelanin intermediate mid: 50 to <75, Eumelanin intermediate high: 75 to <100, and Eumelanin high: ≥100. Melanin index cutoff values were based on measurements from the ColorMeter DSM III from Cortex Technologies. Similarly, the measurements used in the Dadzie study included data collected on the DermaSpectrometer (Cortex Technologies), the ColorMeter DSM II (Cortex Technologies), and the Mexameter-MX18 (Courage+Khazaka Electronics, GmbH), which are related to but not identical to the Melanin Index measurements from the SkinColorCatch®, so a conversion equation was ascertained as described below.

The melanin index of 28 different skin tone swatches from across the PANTONE SkinTone^TM^ Guide (Pantone LLC, Carlstadt, NJ) were measured in triplicate with both the SkinColorCatch® and the ColorMeter DSM II. The average melanin values from each device were graphed and a logarithmic relationship between the two devices was identified (R^2^ = 0.9968). The logarithmic equation (Equation 1) was used to convert the SkinColorCatch® melanin values measured at the forearm to the ColorMeter DSM II scale and Eumelanin Scale-Modified categories, accordingly.

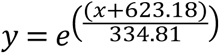

**Equation 1. The logarithmic equation used to convert the SkinColorCatch® melanin values measured at the forearm to the ColorMeter DSM II scale needed for determining the Eumelanin Scale-Modified categories. X represents the Melanin Index measured by the SkinColorCatch® and y represents the melanin value that is used to categorize the Eumelanin Scale-Modified categories.**

#### Thermal Imaging

The FLIR E8-XT and the FLIR ProOne (FLIR Systems, Inc., Wilsonville, OR) thermal cameras were used to collect optical and thermal images of the sacral region throughout the study. The Multi-Spectral Dynamic Imaging (MSX) image setting was used, allowing the visual and thermal image to be seen in a single fusion image.

### Data Collection

The participants’ posterior superior iliac spines (PSIS) were palpated while the participant was standing. A 2” circle was drawn on the skin around each PSIS using an eyeliner pencil with high color contrast to the participant’s skin tone. This circle was traced to ensure a solid and clear marking. The right PSIS was used for the cooling protocol.

The overall flow of the study was conducted according to Figure 1. A control image (prior to any cooling of the skin) was collected with both thermal cameras with the participant side-lying with their hips and knees bent to approximately 90 degrees, legs on top of one another and a pillow placed between their knees (knees stacked, Figure 2) to simulate a clinically relevant and repeatable posture. This generally occurred after the participant had approximately 15 minutes to acclimate to the room while the consent process and demographic survey were completed. Both control images were taken with the camera 50 cm from the PSIS with ambient lighting.

**Figure 1.**
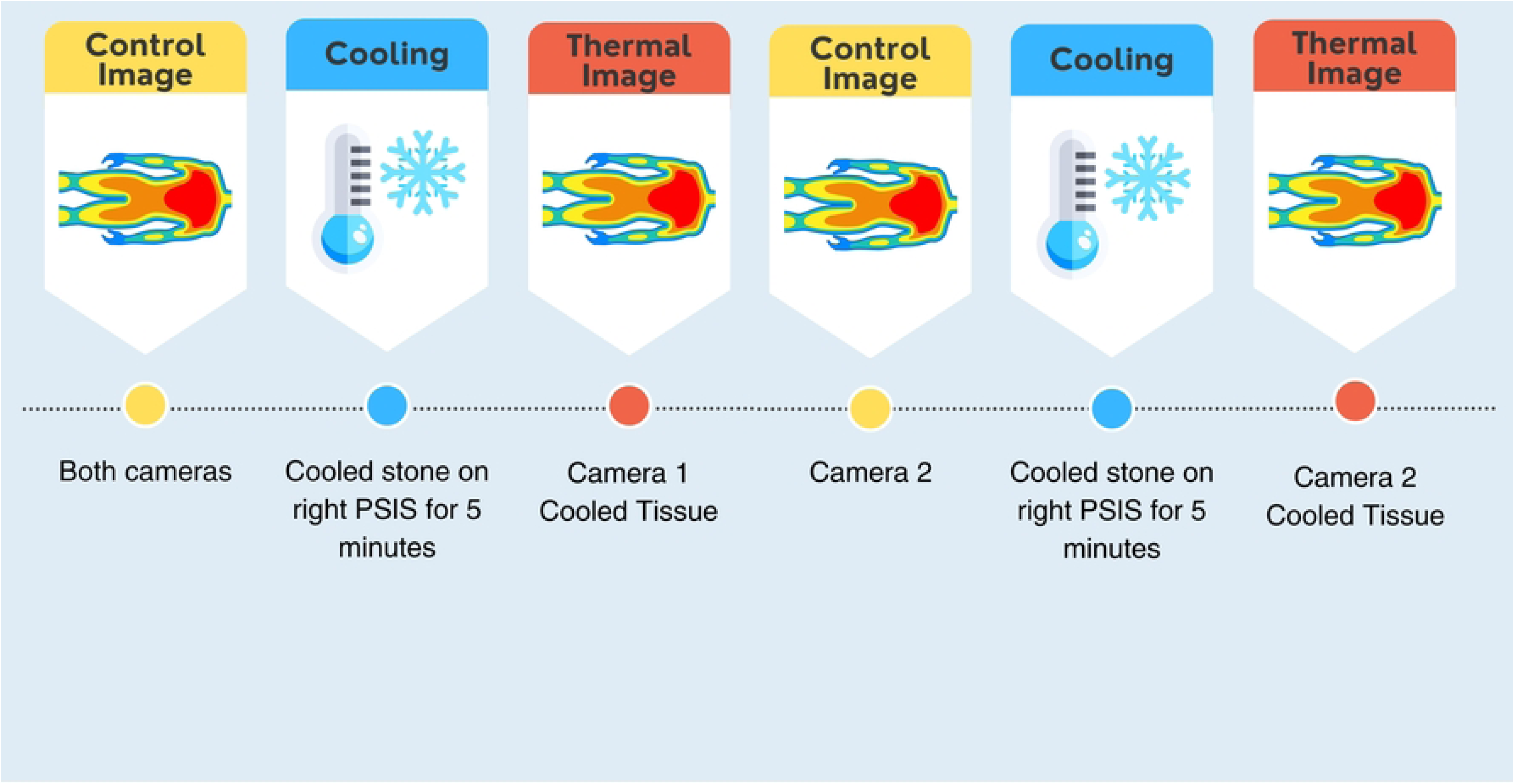
**Study protocol includes two cooling interventions – once for each test camera.**

**Figure 2.**
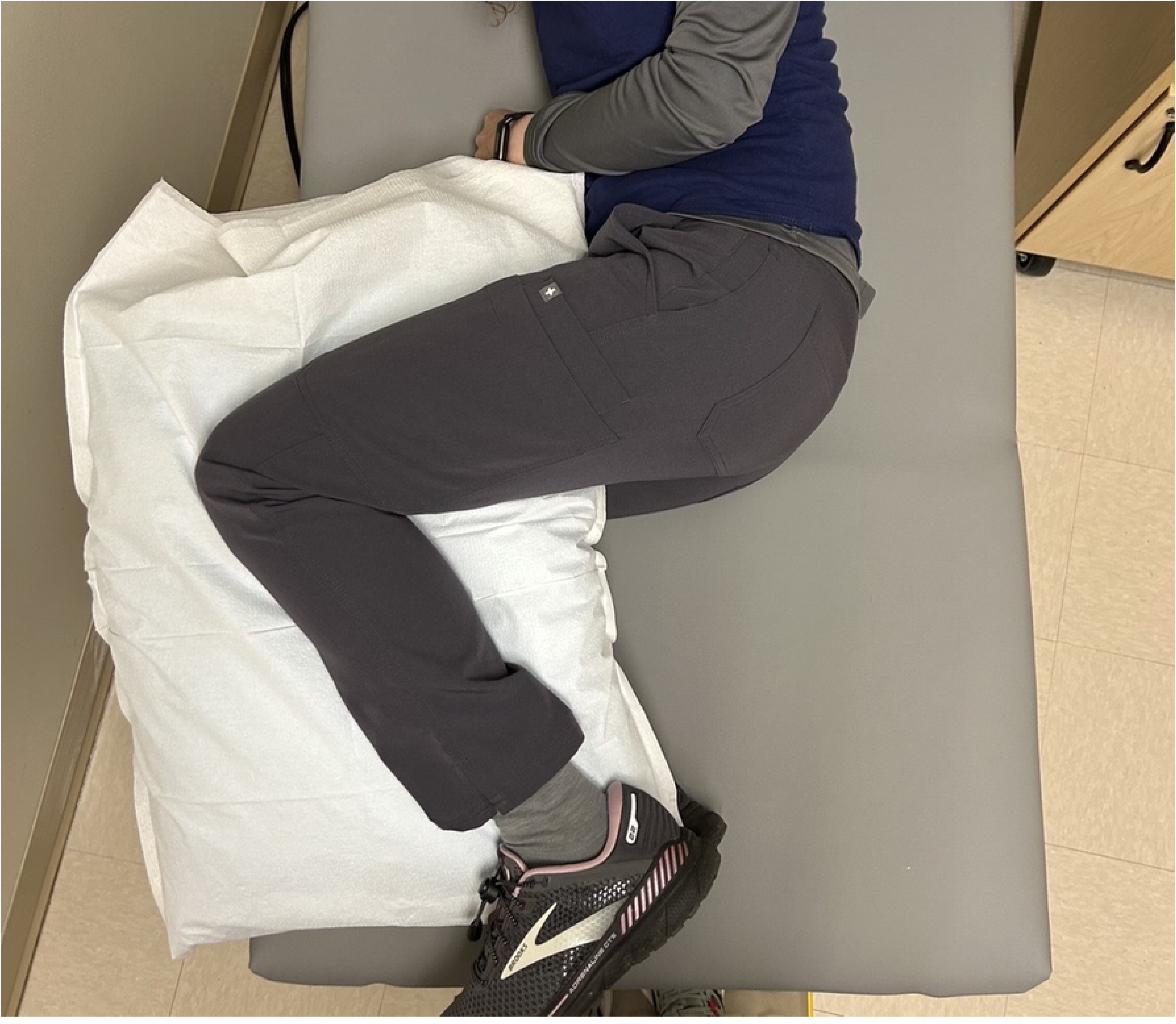
**Reference posture for data collection**

While the participant was on their side, a series of baseline images were collected with the first, randomly selected thermal camera. These images were taken under different combinations of two lighting conditions (ambient/room lighting on and a ring LED light with room lights off), two camera distances (35 and 50 cm), and 3 postures (Table 1) for a total of 12 unique images. The order of the combinations was randomized for each participant. Next, the patient was asked to lay prone, while a stone that had been cooled in a 60-degree Fahrenheit water bath was placed on their right PSIS for 5 minutes. The participant then returned to lying on their side to repeat the same series of 12 imaging conditions with the first camera to capture post-cooling measurements. This ended the first cooling protocol with 24 pre-post paired images, 12 baseline and 12 cooled.

**Table 1.** Conditions for thermography images will consider the variations experienced in a clinical environment.

| Lighting | Distance from PSIS | Patient Position |
| --- | --- | --- |
| Ambient (Room) Lighting | 35 cm | Side-lying with legs on top of one another and pillow between knees (knees stacked) |
| Room lights off, bright LED ring light on | 50 cm <sup>19</sup> | Side lying with forward placement of top knee, creating an increase in flexion of the top hip (knee forward)<br>Side-lying with rearward placement top knee, creating a decrease in flexion of top hip (knee behind) |

After the first cooling protocol, the ROI was given approximately 15 minutes to return to baseline temperature before the cooling protocol was then repeated on the right PSIS. This time images were collected with the second thermal camera. The order of the 12-image series (i.e., the randomized sequence of the combinations of lighting, distance, and patient positioning conditions) was kept the same as with the previous thermal camera. This completed the study visit with a total of 48 pre-post paired cooled images.

### Data Processing

The corner points on the identification card and/or sticker visible in the image were mapped between the optical and thermal images with an affine transformation, aligning the two images.

To process the cooling images, two regions of interest (ROI) were selected (Figure 3). An elliptical region of interest was selected using a custom Python script that matched the outline of the circle drawn on the PSIS. We defined the ROI by placing a minimum of five points along the border of the circle and used these points to generate a best-fit ellipse that encompassed the cooling region.

**Figure 3.**
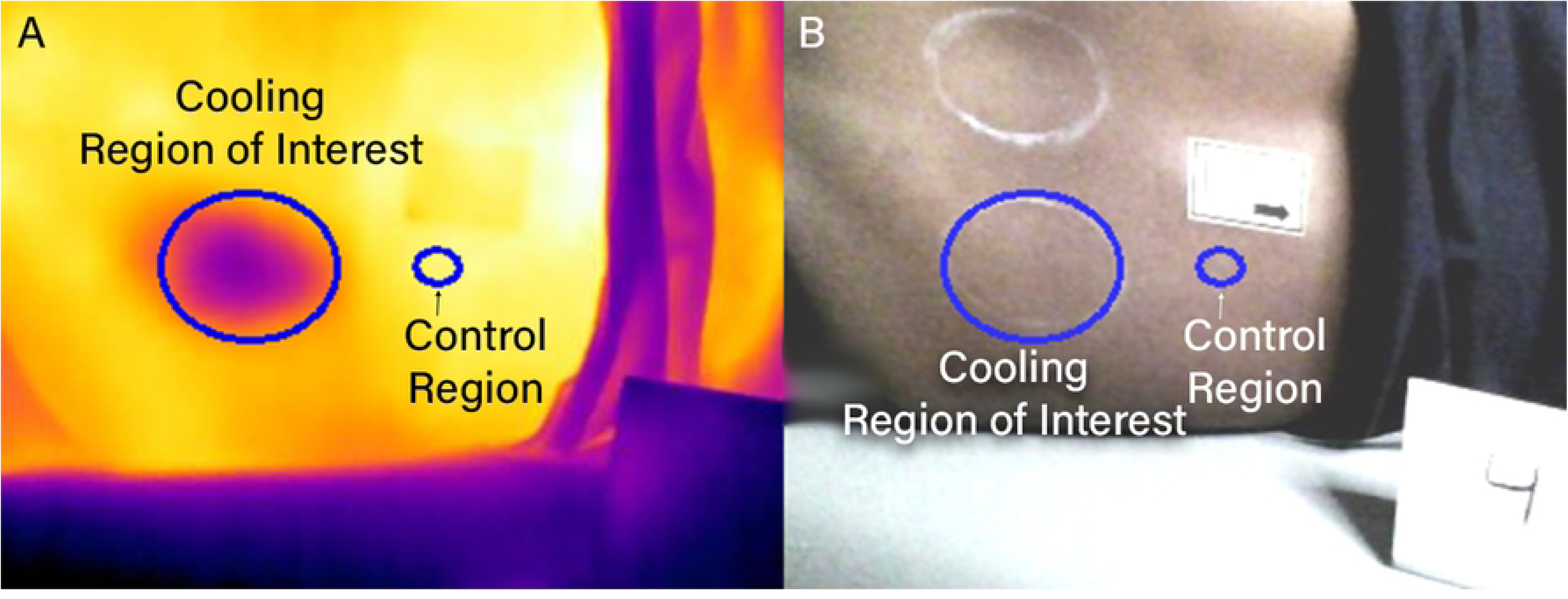
The two regions of interest identified for the cooling protocol are illustrated on the (A) thermal image and (B) optical image following alignment via transformation.

A standardized control region was automatically created as an ellipse scaled proportionally to the cooling ROI, with axes measuring 1/4th the length of the long and short axes of the cooling ROI. This scaling ensured the control region captured a consistent proportion of tissue across participants regardless of anatomical variations. The control region was positioned superior along the x-axis of the image from the center of the original cooling ROI, offset by the width of the ellipse plus the radius of the long axis of the original cooling ROI. On some occasions, this placed the control region overlapping with the sticker or clothing, in which case it was manually relocated to the nearest appropriate location consistent with the originally defined parameters while avoiding any overlap. This standardized approach balanced the need for adequate tissue sampling against the sensitivity of single-point measurements and the challenge of using full-sized control ROIs (which would face inadequate control region availability).

### Data Analysis

Statistically significant differences between baseline and cooling periods within a single region were assessed using a one-sample t-test (Δ = 0). Control and cooling ROI were compared at individual timepoints (baseline or cooling) using paired t-tests. Linear mixed-effects models were used to evaluate the effect of camera type on temperature measurements. A separate linear mixed-effects model, using only data collected with the FLIR E8-XT camera, was conducted to evaluate the effects of distance, participant posture, lighting, and skin tone. Skin tone was analyzed as a continuous variable using melanin index measurements obtained from the forearm using the SkinColorCatch.

## Results

### Participants<colcnt=2>

This study included 35 participants who varied in age, ranging from 18 to 73 years old (Table 2). We recruited 30 participants who had a MST level of 6 or greater, and 5 participants who had a MST level of 5 or lighter. Study participants included more women, and were evenly dispersed between those of normal BMI, overweight and obese. Due to our enrollment strategy of including 30 participants who had a MST level of 6 or greater, most participants were in the Intermediate Mid Modified Eumelanin Skin Tone Category (40).

**Table 2.**
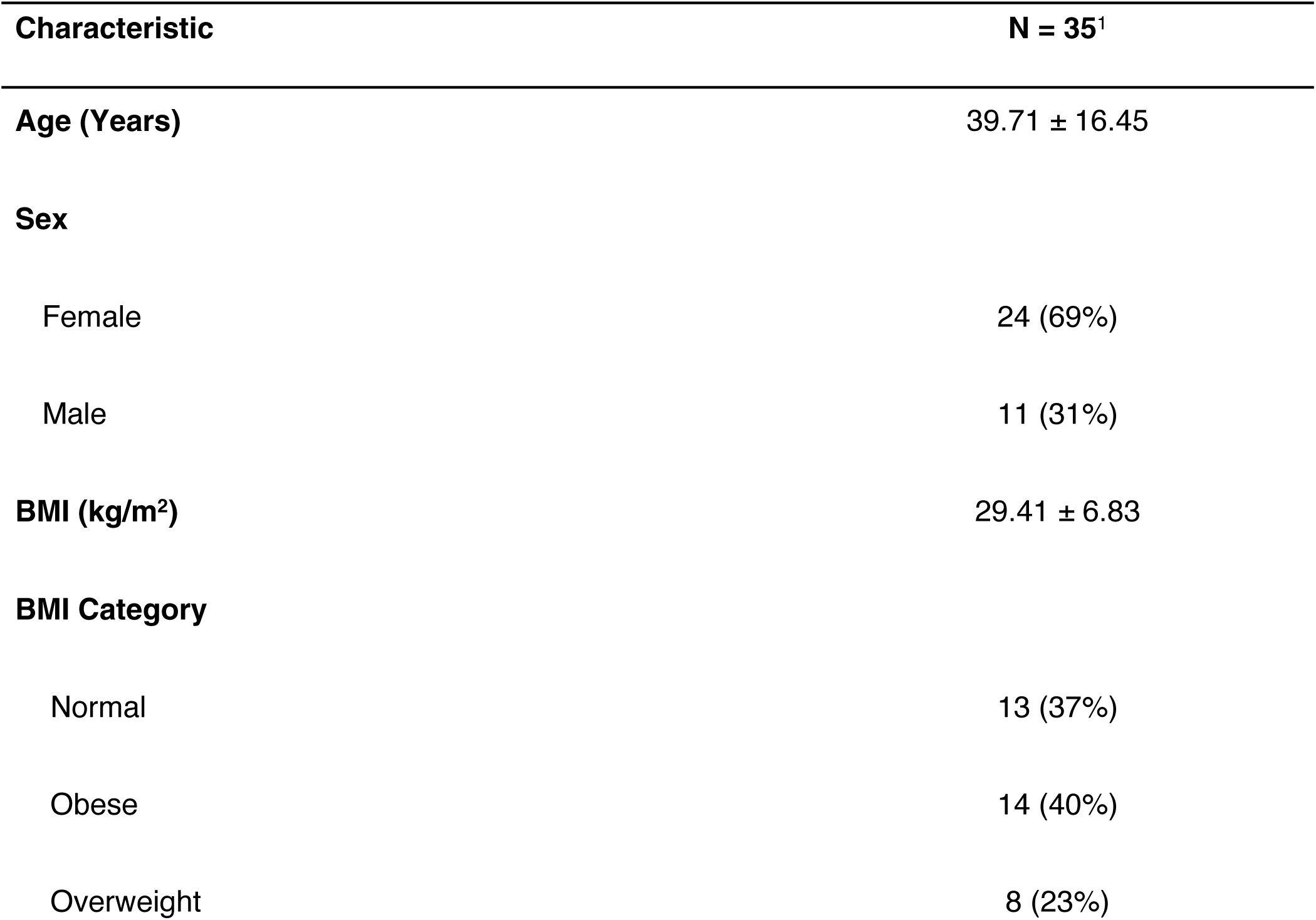
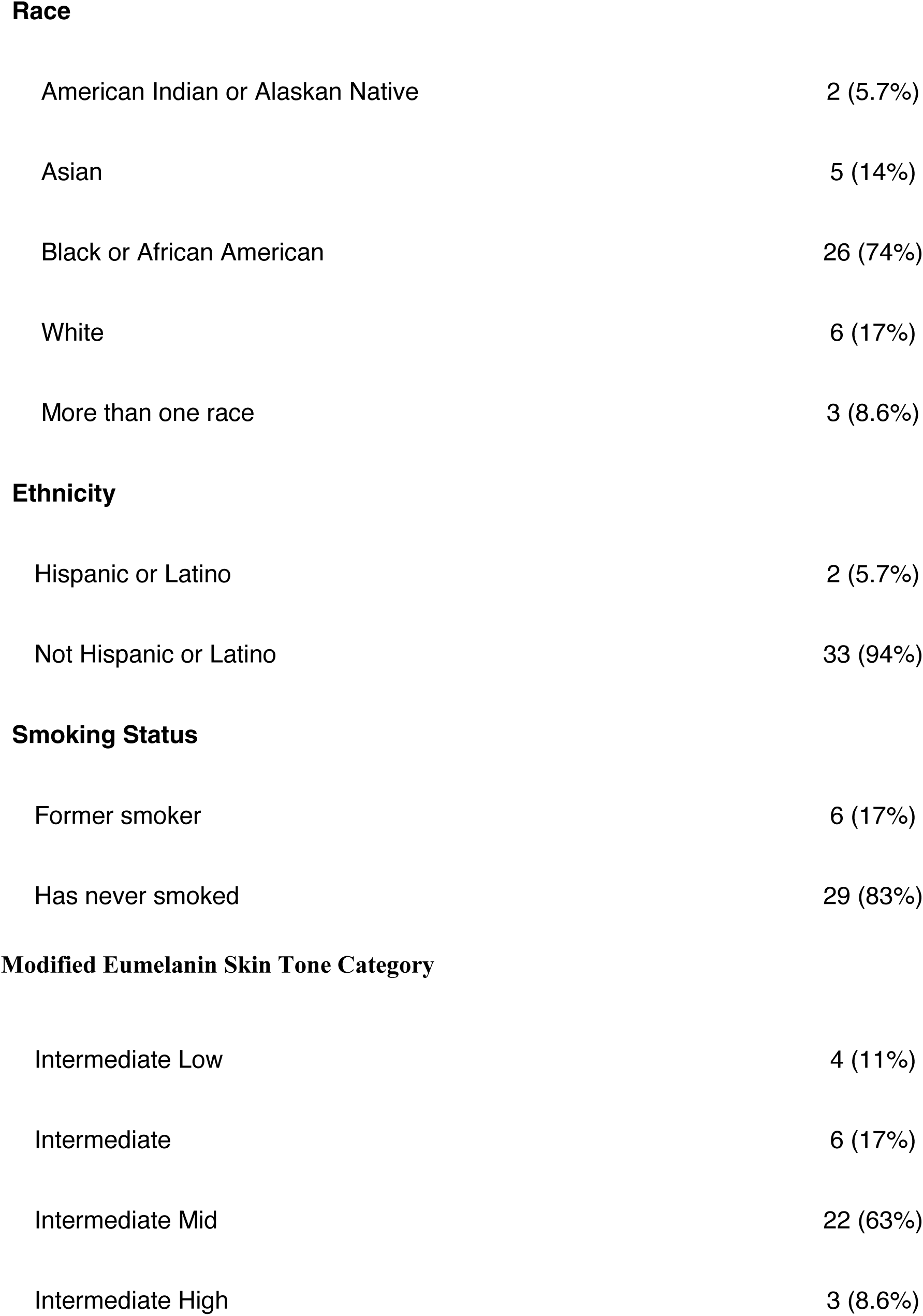
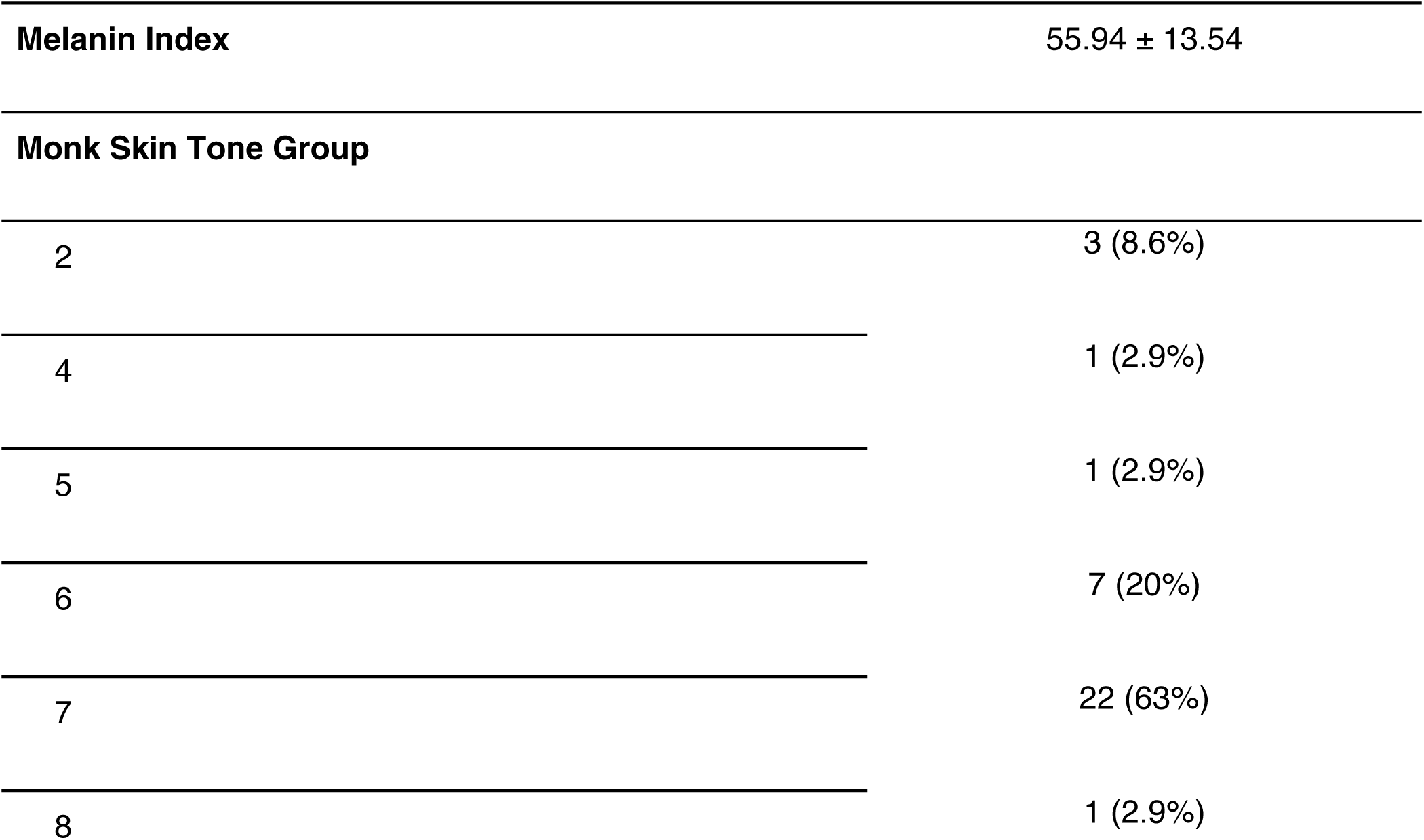
Participant characteristics.

| Characteristic | N = 35<sup>1</sup> |
| --- | --- |
| **Age (Years)** | 39.71 ± 16.45 |
| **Sex** |  |
| Female | 24 (69%) |
| Male | 11 (31%) |
| **BMI (kg/m<sup>2</sup>)** | 29.41 ± 6.83 |
| **BMI Category** |  |
| Normal | 13 (37%) |
| Obese | 14 (40%) |
| Overweight | 8 (23%) |

| **Characteristic** | **N = 35<sup>1</sup>** |
| --- | --- |
| **Race** |  |
| American Indian or Alaskan Native | 2 (5.7%) |
| Asian | 5 (14%) |
| Black or African American | 26 (74%) |
| White | 6 (17%) |
| More than one race | 3 (8.6%) |
| **Ethnicity** |  |
| Hispanic or Latino | 2 (5.7%) |
| Not Hispanic or Latino | 33 (94%) |
| **Smoking Status** |  |
| Former smoker | 6 (17%) |
| Has never smoked | 29 (83%) |
| **Modified Eumelanin Skin Tone Category** |  |
| Intermediate Low | 4 (11%) |
| Intermediate | 6 (17%) |
| Intermediate Mid | 22 (63%) |
| Intermediate High | 3 (8.6%) |
| Melanin Index | 55.94 ± 13.54 |
| Monk Skin Tone Group |  |
| 2 | 3 (8.6%) |
| 4 | 1 (2.9%) |
| 5 | 1 (2.9%) |
| 6 | 7 (20%) |
| 7 | 22 (63%) |
| 8 | 1 (2.9%) |

### Temperature Change after Cooling

Temperature in the ROI was 30.8°C ± 2.0°C prior to cooling and use of the cooling stone resulted in 3.2° C ± 1.7° C of cooling (Table 3). Temperatures in both the ROI and the control region decreased significantly following cooling, but the average cooling in the control region was -0.2 ° C ± 1.3° C from baseline.

**Table 3.** Temperature responses during baseline and cooling in both regions of interest.

| **Median Temperature in Region** | **Baseline**<br>N = 840<sup>1</sup> | **Cooled**<br>N = 839<sup>1</sup> | **Baseline – Cooled**<br>N = 839<sup>1</sup> |
| --- | --- | --- | --- |
| Cooling Region of Interest (deg C) | 30.8 ± 2.0 | 27.5 ± 1.5 | -3.2 ± 1.7* |
| Control Region | 31.1 ± 1.8 | 31.2 ± 1.4 | -0.2 ± 1.3* |
| Cooling Region of Interest – Control Region | -0.3 ± 0.6** | -3.7 ± 1.2** |  |

<sup>1</sup>Mean ± SD
\* p<0.001 one sample t-test, Δ=0
\*\* p<0.001 paired t-test comparing the cooling ROI to the control ROI during baseline or cooling

### Differences across Environmental and Technical Factors

A linear mixed-effects model revealed significant differences between cameras and their measurement of temperature changes (Figure 4). Under baseline conditions, measurements from the ProOne camera were 2.43°C lower than those from the FLIR E8-XT (p < 0.001). With the FLIR E8-XT camera, cooling reduced temperatures from baseline by 4.19°C (p < 0.001). However, a significant interaction between cooling and camera type (β = 1.89, p < 0.001) indicated that the difference in the measured temperature change between cameras was 1.89°C smaller when measured with the ProOne camera. When analyzing differences between ROI and control regions, which is more clinically relevant, there was no significant difference between cameras at baseline (β = -0.08°C, p = 0.113). After cooling, the ROI temperature decreased 3.95°C more than the control region when measured with the FLIR E8-XT (p < 0.001). A significant interaction term (β = 1.04°C, p < 0.001) indicated that this difference was 1.04°C smaller when measured with the ProOne camera. However, both devices still observed significant temperature changes across participants.

**Figure 4.**
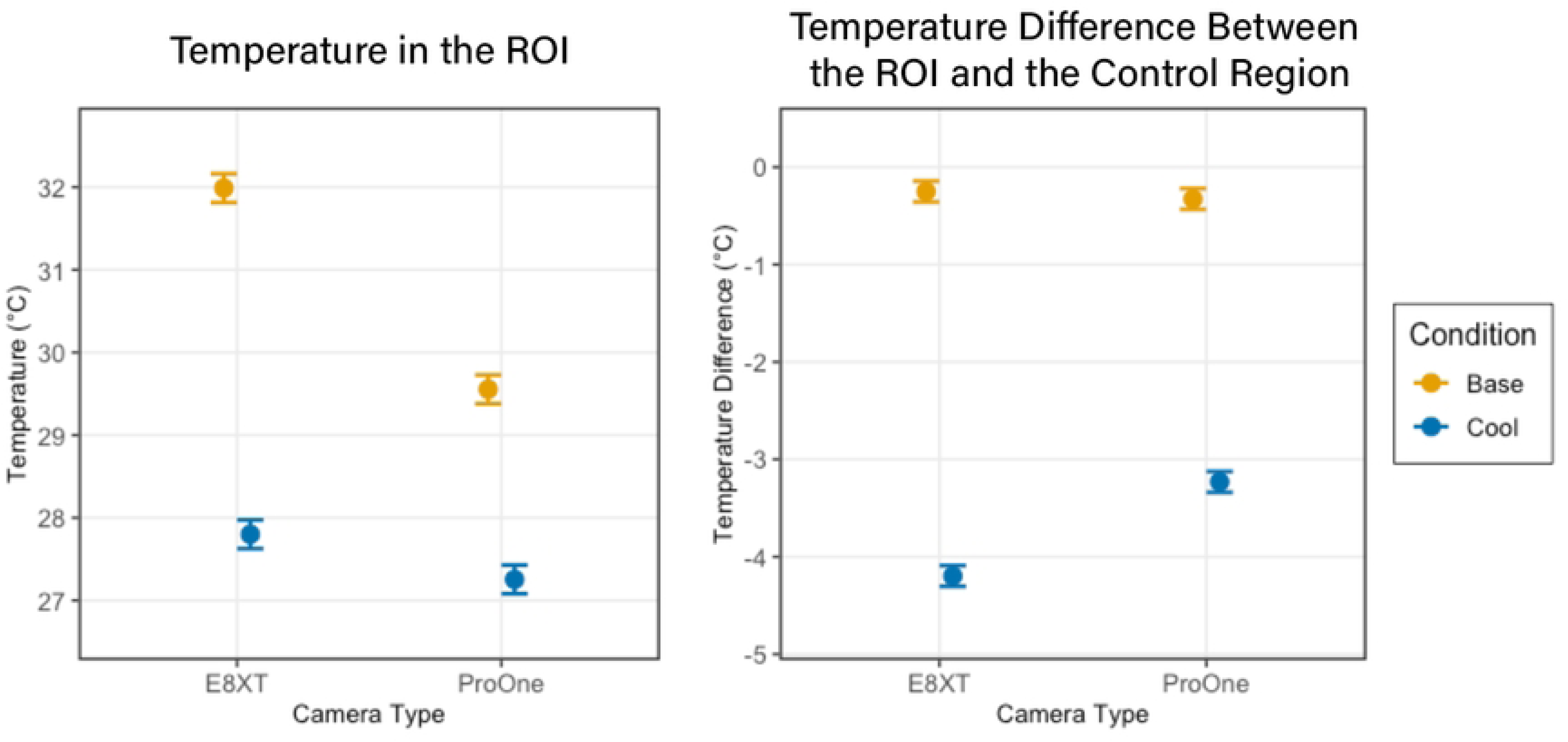
There was a significant impact of cooling and camera on the absolute temperature in the ROI (Left), whereas the temperature difference between the ROI and Control Region showed camera-dependent variation only in the cooling condition (Right).

Further analysis was all run using only data collected on the E8-XT camera. Distance had small but variable effects on temperature measurements. At 50 cm, ROI temperatures were 0.11°C cooler than at 35 cm during baseline, though this difference was smaller during cooling (interaction β = 0.20°C, p = 0.045). Control regions showed a similar but non-significant trend to be cooler at 50 cm (β = -0.07°C, p = 0.177). When analyzing the more clinically relevant metric, the difference between ROI and control regions, measurements at 50 cm were not significantly different from those at 35 cm (β = 0.08°C, p = 0.122).

Linear mixed effects models confirmed a significant temperature reduction during cooling compared to baseline. However, neither patient positioning nor lighting conditions significantly affected the measurement of temperature change, whether assessed as absolute temperature in the cooling region or as the temperature difference between cooling and control regions.

### Differences across Skin Tone

Linear mixed effects modeling revealed a significant interaction between melanin and cooling (p = 0.001), indicating that the thermal camera’s measurement of the temperature change varied with skin tone (Figure 5). While cooling produced an overall temperature reduction in the difference between the ROI and the control region (estimate = -3.24°C, p < 0.001), this difference was slightly increased in participants with higher melanin levels, as evidenced by the negative interaction term (estimate = -0.013°C per color unit, p = 0.001). Because different skin tone categories are separated by 25 units, this would represent a measured difference of 0.325°C per skin tone category, or a difference of 0.975°C between the Intermediate Low and the Intermediate High skin tone categories.

**Figure 5.**
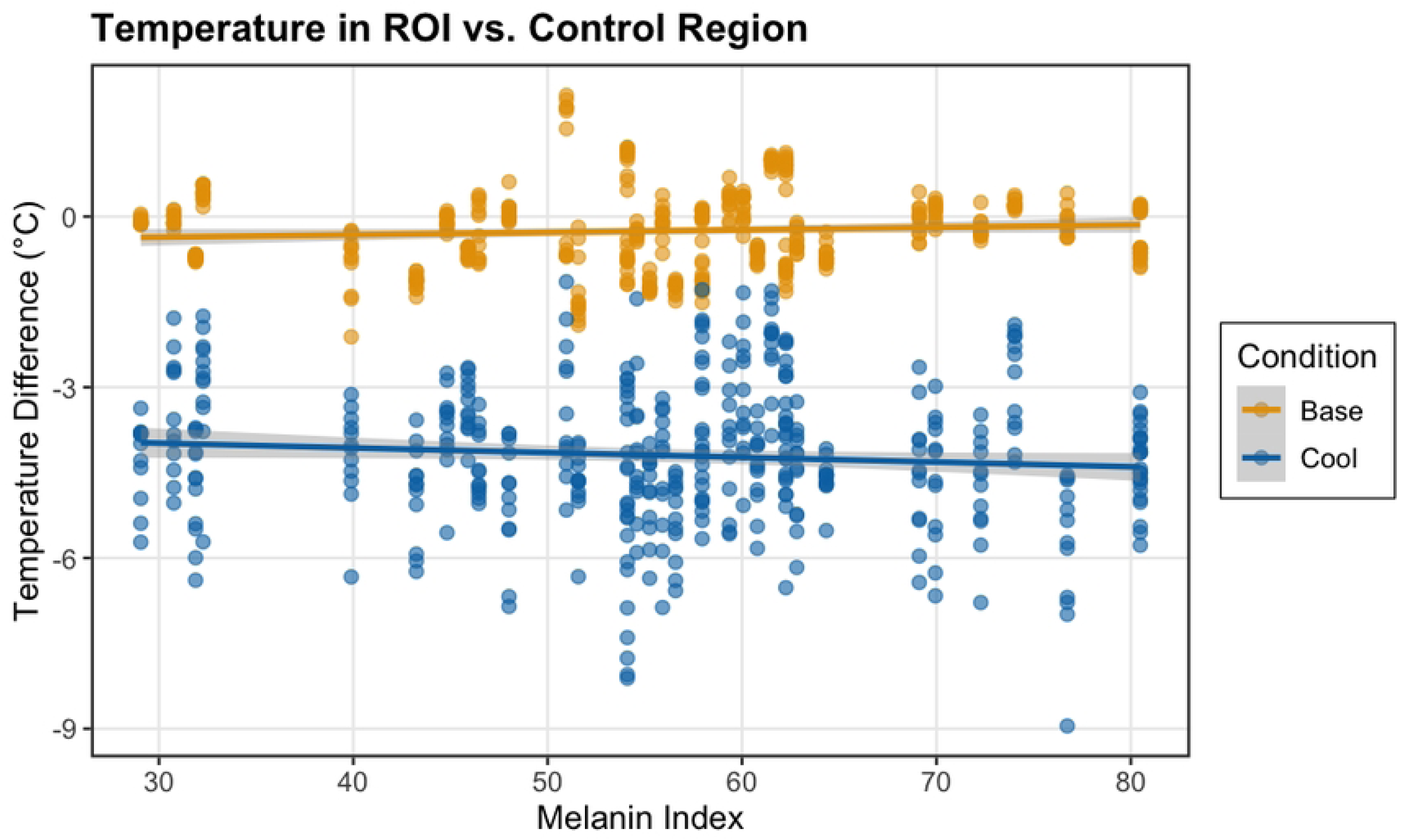
Measured temperature difference between the ROI and control region at baseline and after cooling varies according to melanin index.

## Discussion

### The impact of environmental and technical factors on thermal measurements and temperature change

Thermal imaging has been tested in highly controlled conditions and implemented within clinical bundles, but the impact of environmental conditions and skin tone have not been reported outside of manufactures’ 510(k) reporting. (43) The present study showed that the FLIR E8-XT thermal camera was not sensitive to lighting or posture. Absolute temperature measurements varied with distance, but the temperature difference between the ROI and control regions remained constant across distances. However, maintaining consistent measurement distances remains important for precise temperature assessment. From a clinical perspective, choosing a control region at a similar distance to the device, which is feasible if the skin surface is perpendicular to the thermal camera, should mitigate the impact of the distance effect. Additionally, varying distances from the camera within a single image, as would occur if the image is not taken perpendicular to the skin surface or is taken over a highly contoured area, could create more significant artifacts and therefore presents a greater concern for clinical interpretation.

In this study, we tested two camera models with substantially different specifications and costs. Our results revealed that these cameras reported temperature changes after cooling with different sensitivities, with discrepancies exceeding 1°C. This finding suggests that equivalency cannot be assumed across all thermographic imaging products. Our results align with what might be expected given the technological diversity documented by Baron et al., who identified six different technologies implemented across eight studies. (8) Among the three devices with available information, measurement precision varied considerably (±2% to ±10%), as did temperature ranges (-20°C to 250°C, -10°C to 140°C, and -20°C to 400°C), while no specifications were available for the remaining three devices. This technological heterogeneity may contribute to the wide variety of temperature thresholds used and identified in the literature to determine tissue damage, ranging anywhere from 0.1°C (5,44), 1°C (45), 1.2°C (1), 1.5°C (46), and 2.2°C (31). Although this list is not exhaustive, it highlights significant gaps in our understanding and the lack of standardization in accepted evidence. Based on these findings, researchers and clinicians should exercise caution when generalizing results and recommended thresholds across different thermographic devices without first considering their technical specifications.

### The impact of skin tone on thermal measurements and temperature change

Variations in temperature were observed across different skin tones, with participants with higher melanin content measuring colder after the same cooling protocol. This pattern resembles previous findings with infrared thermometers, where lower temperature readings in African-American patients led to the under detection of fevers, though the magnitude of difference was considerably different between studies. (34) Whether this variation stems from actual physiological differences in thermodynamic response to our intervention or from measurement artifacts related to thermal imaging remains undetermined. Nevertheless, two key conclusions emerged: First, thermal imaging successfully detected temperature changes across all skin tones, confirming its utility for detecting cooling in individuals with darker skin. This was anticipated based on the constant emissivity across skin pigmentation. (28) Second, our findings emphasize the necessity for rigorous clinical validation across diverse skin pigmentations. Since our research also aims to detect temperature increases due to inflammation, which may register as smaller changes in individuals with higher melanin content, developing melanin-specific detection thresholds could be warranted if measurement sensitivity varies with skin pigmentation.

## Conclusion

In conclusion, this study provides valuable insights into the reliability and limitations of thermal imaging across diverse skin tones. Our findings demonstrate that while distance had a small but measurable effect on absolute temperature measurements, this can be effectively mitigated through perpendicular imaging and selecting control regions at consistent distances. The observed discrepancies exceeding 1°C between different thermal cameras indicate that these technologies cannot be used interchangeably, particularly when establishing detection and healing thresholds. Most notably, our discovery that participants with higher melanin content exhibited lower temperature readings following identical cooling protocols suggests that skin pigmentation directly impacts thermal imaging assessment. These results emphasize the potential need for melanin-specific detection thresholds to ensure equitable diagnostic accuracy across diverse populations. Future research should focus on developing standardized protocols that account for both technological variations and skin tone diversity to improve the clinical utility of thermal imaging.

## Data Availability

URLs/accession numbers/DOIs will be available only after acceptance of the manuscript.

## Acknowledgements

This study was supported in part by the National Center for Advancing Translational Sciences of the National Institutes of Health under Award number UL1TR002378. The content is solely the responsibility of the authors and does not necessarily represent the official views of the National Institutes of Health.

